# Sense of coherence and self-care in people with diabetes: systematic review

**DOI:** 10.1101/2025.01.16.25320434

**Authors:** María del Carmen Vega-Martínez, Catalina López-Martínez, Rafael del-Pino-Casado

## Abstract

**Introduction:** Diabetes mellitus is one of the diseases with the greatest social and health impact. Self-care in people with diabetes requires constant physical and emotional effort, which is one of the barriers to adherence to the care plan. Sense of coherence can play an important role in self-care.

**Aims:** To examine the relationship between sense of coherence and self-care in individuals with diabetes mellitus.

**Method:** A systematic review with narrative synthesis for all included studies and with meta-analysis in those with adequate statistical data. We searched PubMed, CINAHL, PsychInfo and Scopus up to July 2024. We included original studies that reported effect sizes on the association between sense of coherence and self-management of care plans in individuals with diabetes. Methodological quality of the studies was independently assessed by two researchers by evaluating the risk of selection bias, classification bias and confounding bias. Meta-analysis was performed using a random-effects model and various sensitivity and subgroup analyses were performed to assess the robustness of the meta-analysis result.

**Results:** Thirteen articles were included in the review (six were included in the meta-analysis), whose participants (9,800) were people with type 1 or 2 diabetes. In the studies analyzing the relationship between sense of coherence and adherence to self-care (diet, exercise, and medication), a positive association was observed between these variables. In addition, sense of coherence may have a protective effect against negative perceptions of self-care. In the meta-analysis, a positive and moderate association between sense of coherence and adherence to self-care was found, with no heterogeneity and possible publication bias that does not affect the results of the meta-analysis.

**Limitations:** Small number of studies; all were cross-sectional.

**Conclusions:** Sense of coherence may play a relevant role in improving adherence to the self-care plan in people with type 1 or 2 diabetes.

**PROSPERO id:** CRD42023390705

## Introduction

In 2021, the International Diabetes Federation reported that 536.6 million people worldwide were living with diabetes, with Spain ranking as the second country in Europe with the highest diabetes prevalence. Additionally, 6.7 million deaths globally were attribute to this disease [1]. Diabetes mellitus (DM) is one of the diseases with the greatest social and health impact, not only due to its high prevalence, but also because of the chronic complications it produces and its high mortality rate. The prevalence of the different chronic complications varies depending on the type of DM, duration of the disease and degree of metabolic control [2]. It is expected that by 2045 there will be around 783.2 million people with DM [1].

In recent years, self-care management has emerged as a powerful ally in disease management, especially in chronic diseases, due to the effect on health outcomes [3]. Daily self-management of DM involves actions that lead to the balance blood glucose fluctuations through the administration of exogenous insulin, in the cases of insulin-dependent people, food intake, physical activity, and self-monitoring, mainly [4].

On a personal level, self-care requires constant physical and emotional effort, which seems to be one of the major impediments to adherence to the care plan by people with DM [5]. Consequently, self-care can be perceived as frustrating, overwhelming, and even a personal overload [6]. In addition, DM-related stress has been studied to understand its relationship with glycemic control. In this sense, a clear relationship has been identified between stress and glycemic control, obtaining worse results in patients with stress or a loss of glycemic control due to minimization of self-care activities, which negatively affects the quality of life of people with DM [7].

The fear of hypoglycemia has been described as another event that produces stress in people with DM, especially if they have had experiences in the past, it can generate anxiety and therefore worse glycemic control [8]. These behaviors increase the risk of complications associated with hyperglycemia and reduce the effectiveness of care plans for optimal glycemic control [9].

Although healthy lifestyles and self-care management are strongly recommended, the implementation of the recommendations is sometimes unsatisfactory [10]. On the other hand, the data supporting the efficacy of self-care in improving outcomes for patients with specific chronic diseases are still surprisingly weak [11].

An important concept in relation to health is the sense of coherence (SOC), which leads people towards optimal outcomes under difficult conditions [12]. Antonovsky defines SOC as a global orientation that expresses the extent to which one has a pervasive, enduring though dynamic feeling of confidence that one’s internal and external environments are predictable and that there is a high probability that things will work out as well as can reasonably be expected [13].

Antonovsky’s salutogenic theory emphasizes the importance of Sense of Coherence (SOC)—comprising comprehensibility, manageability, and meaningfulness—in the management of chronic conditions such as diabetes. A strong SOC may enable individuals to better understand their condition, access necessary resources and find purpose in managing their health [12] This, in turn, might fosters improved self-care behaviors, such as glucose monitoring and treatment adherence, while helping individuals view their illness as manageable and their efforts as meaningful [14,15], ultimately enhancing decision-making in diabetes management.

In recent years, the study on the relationship between SOC and self-care issues in people with DM, such as good dietary choices, exercise, pharmacological treatment control, stress and disease burden has gained importance. Authors have shown that SOC is related to better health outcomes [16–19]. Despite this, there is still limited literature providing evidence on the relationship between SOC and self-care plan. Some studies indicate that SOC is related to better adherence and positive perception of the self-care plan [20–22], while others do not report these results [19,23,24]. Therefore, the aim of this research is to provide a systematic and up to date synthesis of the available evidence of the relationship between sense of coherence and self-care in individuals with diabetes mellitus.

## Materials and Methods

### Design

A systematic review with meta-analysis was conducted in accordance with the guidelines of the Preferred Reporting Items for Systematic reviews and Meta-Analyses (PRISMA) [25] to ensure a logical and research-validated approach. We pre-registered our review in the International Prospective Register of Systematic Reviews (PROSPERO id: CRD42023390705; Available in: https://www.crd.york.ac.uk/prospero/).

### Search strategy

A literature search was conducted in the main health science databases (PubMed, PsycINFO, CINAHL, and Scopus) from the first year included in each database until July 2024. To build the search strategy, we developed the following search question: What is the relationship between the sense of coherence and self-care management in people with diabetes? From this question, we chose the following terms for the search: diabetes, sense of coherence, adherence to treatment, diet, physical activity, medication, self-care, self-management and care plan, which we combined with the Boolean operators “AND” and “OR”, and parenthesis (see S1 Appendix). No additional filters for language, time, or study design were used. Grey literature and citation searching (backward and forward) were also used in the search.

### Inclusion criteria

Inclusion criteria were as follows: (1) original studies (2) reporting on the relationship between SOC and self-care or self-management of the care plan (3) in people with diabetes (type 1 [T1D], type 2 [T2D] and gestational diabetes [GDM]), (4) and reporting the correlation coefficient of the relationship or another statistical value compatible with this correlation coefficient. Two reviewers (MCVM and RDPC) independently reviewed and selected studies. Discrepancies were resolved by consensus.

### Data extraction

Using a standardized data collection form, two investigators (MCVM and RDPC) independently extracted data regarding: design, sample size, mean age and range, percentage of women, type of diabetes, instruments to measure SOC and care plan management, type of pharmacological treatment, and self-care referred to in the study. Disagreements were resolved by consensus.

The effect measure used for the calculation of the combined estimates was the correlation coefficient.

### Quality assessment of included studies

The methodological quality assessment (risk of bias) of the included articles was conducted based on recommendations of Boyle [26] and Viswanathan et al. [27], using the following criteria: (1) for selection bias: type of sampling (probabilistic/non-probabilistic) and for longitudinal studies, attrition rate (no more than 20%); (2) for classification bias: reliability and validity of measures: content validity and internal consistency and 3) for confounding bias: control or adjustment of potential confounding factors. Two review authors (MCVM and RDPC) independently assessed quality of the included studies. Disagreements were resolved by consensus.

For the analysis of confounding bias, it was decided to select the variables age, sex, and type of diabetes, since factors influencing medication adherence include advanced age, female sex [28] and the type of diabetes the person suffers. Regarding this last aspect, it is estimated that non-adherence to therapeutic treatment is around 50-60% in T2D [29] and more variable in T1D, ranging from 12 to 60% [30].

We consider potential confounders to be controlled when group allocation between groups or matching was appropriate (e.g., through stratification, matching or propensity scores) or confounding factors are controlled in the design and/or analysis (e.g., through matching, stratification, interaction terms, multivariate analysis, or other statistical adjustment as instrumental variables) [31]. In cases of statistical adjustment, we consider that there is no confounding bias when the variance of the point estimate is less than 10% [32].

### Certainty assessment

The quality of the meta-analysis results was evaluated by analyzing inconsistency, imprecision, and publication bias in accordance with the Grading of Recommendation, Assessment, Development, and Evaluation (GRADE) guidelines s [33]. Inconsistency was assessed through heterogeneity of findings in individual studies and imprecision through the number of included studies (large>10 studies; moderate: 5-10 studies; small <5 studies) and median sample size (high>300 participants; intermediate: 100-300 participants; and low < 100 participants) [34]. Finally, publication bias was assessed by analyzing the funnel chart and relevant statistical tests.

### Analysis

We developed a meta-analysis of correlation coefficients using a random-effects model, according to the recommendations of Cooper et al. [35], to estimate combined effects and their 95% confidence intervals (CIs). We also used effect sizes convertible into correlation coefficients, such as standardized mean differences and odds ratios. Conversions were performed by the meta-analysis statistical program.

For the heterogeneity analysis, the Q test was performed, supplemented by calculating the degree of inconsistency (I^2^) of Higgins et al. [36] and the prediction interval [37].

Publication bias was assessed using the funnel plot, the Egger’s test [38] (low risk of publication bias if p>0.1) and the Trim and Fill method [39] (calculation of the combined effect in a situation of absence of publication bias).

A sensitivity analysis was performed to assess the robustness of the meta-analysis results by calculating the combined effect removing one study each time (leave one out method) [35] and observing the percentage of variation with respect to the original combined effect.

We used subgroup analyses to evaluate the possible influence on the meta-analysis results of methodological quality (whether or not you control for selection, classification and confounding bias), study design (cross-sectional or longitudinal) and the type of DM.

The calculations were performed using the statistical program Comprehensive Meta-analysis 3.3 (Biostat, Englewood, NJ, USA).

## Results

### Description of search results

A total of 402 articles were retrieved from the search. After the removing duplicate articles, 316 articles were reviewed by title and abstract. Of these, 251 articles were excluded because they were not relevant. A total of 65 full-text articles were reviewed and 50 were excluded because they did not meet inclusion criteria and 15 articles were selected for quality assessment. One of these 15 articles was excluded because the journal where it was published rejected it [40]. Finally, 14 articles were included in this review [18–22,41–49]. The flowchart shows the search and selection process (Fig 1).

**Fig 1.**
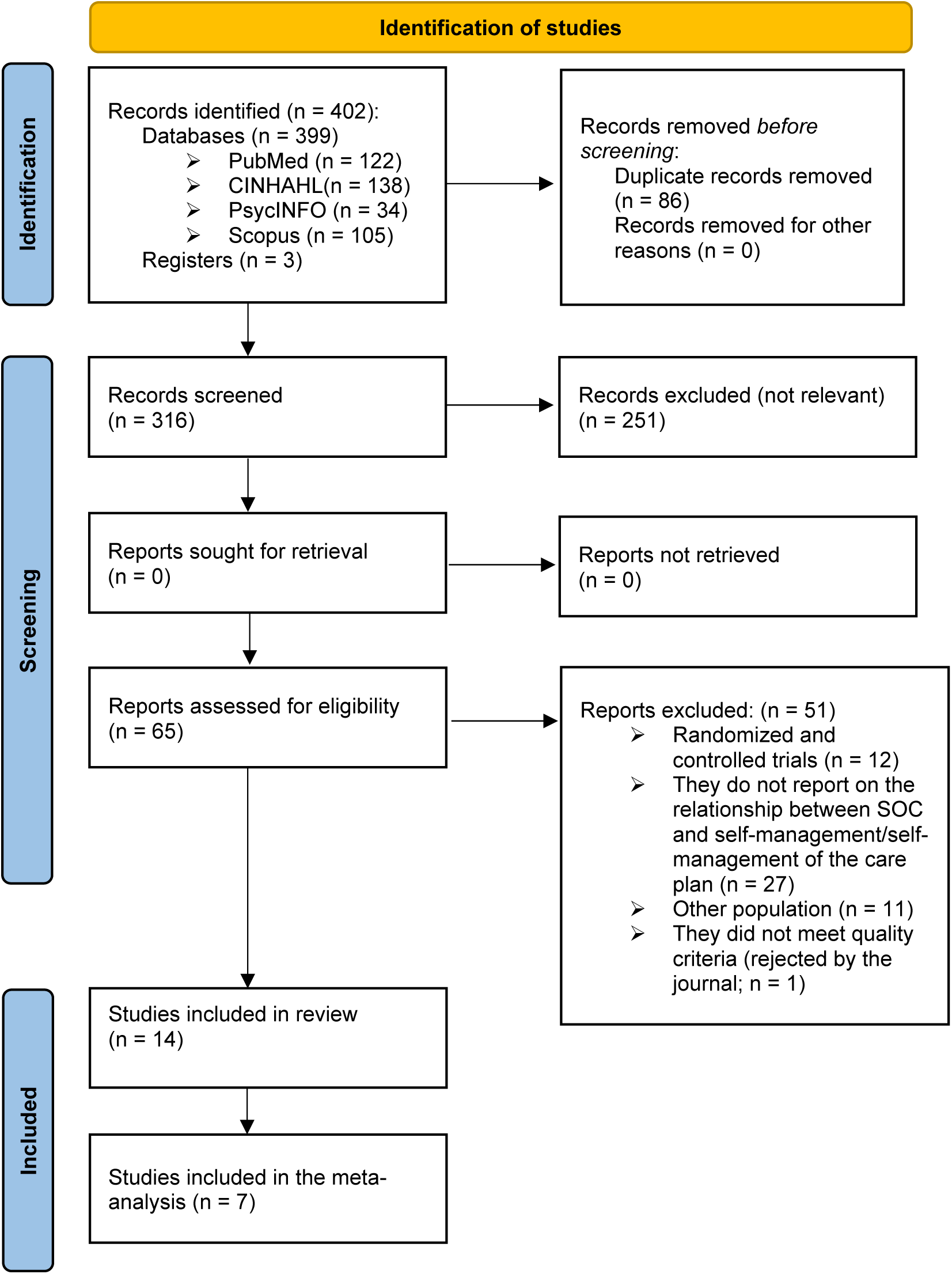
Flow chart of the study selection process.

### Description of included studies

The main characteristics of the studies analyzed are shown in **Table 1**. The total population included was 9,800 adults with T1D [47,48], T2D [18–22,41,42,45,46,49] and both types of diabetes [43,44]. The age range was 18-83 years. The studies examined the relationship between SOC and self-management of the care plan, finding two dimensions: adherence to self-care [18,21,22,41,43–49] and perception of self-care [19,20,42]. Among those analyzing adherence to self-care, we found studies that analyzed adherence to all self-care activities combined [18,22,43–46] and others that focused on specific self-care (diet, exercise or medication) [21,41,47–49]. All included studies were cross-sectional descriptive.

**Table 1.** Characteristics of the studies included in the review.

| Author/year | N | Study design | Mean age (standard deviation) and [range] | % women | Type of DM | SOC Instrument | Type of drug treatment | Outcomes | Measuring instrument |
| --- | --- | --- | --- | --- | --- | --- | --- | --- | --- |
| Ahola 2010 [47] | 1264 | CS | 45 (12) [33-57] | 56 | T1D | SOC-13 | Insulin | Doctor and nurse visits | Ad hoc |
| Ahola 2012 [48] | 1104 | CS | 44 (12) [33-53] | 56.0 | T1D | Soc-13 | Not specified | Diet | Diet questionnaire |
|  |  |  |  |  |  |  |  | Exercise | Leisure Time Physical Activity (LTPA) questionnaire |
| Cohen 2004 [44] | 67 | CS | 52.6 (13) [36-65] | 37.3 | T1D and T2D | SOC-29 | ADO and insulin | Self-care | Adherence to Self-care Behaviors questionnaire from the Self-care Inventory |
| He 2006 [42] | 202 | CS | 62.0 (9) [53-71] | 43.6 | T2D | C-SOC 13 | ADO and insulin | Diabetes-specific stress | Diabetes-specific Stress Perceptions (DSSP) |
| Kordbagheri 2024 [46] | 412 | CS | 58,2 (6,4) [51.8-64.5] | 34.8 | T2D | SOC-29 | Not specified | Self-care | Adherence questionnaire |
| Koponen 2017 [41] | 2866 | CS | 63.0 [27-75] | 43 | T2D | SOC-13 | ADO, insulin or only self-care | Diet and exercise | Sucess in Wieight Management (SWM) |
| Koponen 2019 [21] | 2860 | CS | 63,0 [27-75] | 44.1 | T2D | SOC -13 | ADO, insulin and only self-care | Self-care | Perceived Competence for Diabetes Scale (PCS) |
|  |  |  |  |  |  |  |  | Intake of fruits and vegetables | Fruits, Vegetables and Berries Intake (FVBI) |
| Lei 2018 [45] | 195 | CS (RCT Baseline) | 58.0 (13.2) [22-83] | 45.1 | T2D | SOC-13 | ADO, insulin and only self-care | Self-care | Summary of Diabetes Self-Care Activities Measure (SDSCA) |
| Lin 2009 [18] | 120 | CS | 59.0 [18.82] | Not specified | T2D | SOC-13 | Not specified | Self-care | Diabetes Self-care Behaviour Scale (DSCS) |
| Marquez-Palacios 2021 [22] | 220 | CS | 56.1 (10.4) [20-69] | 74.5 | T2D | SOC-13 | Not specified | Self-care | Summary of Diabetes Self-Care Activities Measure (SDSCA) |
| Odajima 2018 [20] | 177 | CS | 57.9 (10.9) [20-75] | 39.5 | T2D | SOC-13 | Not specified | Burden by self-care plan | Problem Areas in Diabetes (PAID) Survey |
| Renosky 2010 [43] | 111 | CS | 56.0 (14,2) [41.8-70.2] | 60.0 | T1D and T2D | SOC-29 | Not specified | Self-care | Summary of Diabetes Self-Care Activities Measure (SDSCA) |
| Shiu 2004 [19] | 72 | CS | 51.6 (11,7) [39.9-63.3] | 61.1 | T2D | SOC-13 | Insulin | Fear of hypoglycemia | Worry Scale |
| Vega-Martínez, López-Martínez and Del-Pino-Casado [49] | 130 | CS | 65.2 (9.8) [18-82] | 36.2 | T2D | SOC-13 | Not specified | Medication | Morisky-Green test |
|  |  |  |  |  |  |  |  | Mediterranean diet | Mediterranean Diet Follow-up Questionnaire (PREDIMED) |
|  |  |  |  |  |  |  |  | Exercise | International Physical Activity Questionnaire (IPAQ) |

Notes: \*p< 0.05, \*\*p< 0.01
Abbreviations: T1D: Type 1 Diabetes Mellitus; T2D: Type 2 Diabetes Mellitus; CS: cross-sectional study; RCT: Randomized Controlled Trial; SOC: Sense of Coherence.

Most studies used Antonovsky’s SOC scale to measure SOC, with 10 studies incorporating the 13-item version [18–22,41,45,47–49], three studies using the 29-item version [43,44,46] and one using the Chinese version (C-SOC) [42], the latter validated by Shiu in 2004 [19].

### Assessment of the methodological quality of the included studies

Of the 14 studies included, eight had probability sampling [21,41–43,45,47,48], one did not specify the type of sampling that was carried out [18], and the rest had non-probability sampling [19,20,22,44,46]. On the other hand, only one study had risk of classification bias [48], while the rest of the studies analyzed reported the adequacy of the validity and reliability of the measures used. Regarding confounding bias, only three studies found to be at low risk of bias [20,21,49]. **Table 2** shows the assessment of the risk of bias in the studies included in the review.

**Table 2.** Evaluation of the methodological quality of the studies included in the meta-analysis.

|  | Selection bias | Classification bias | Confounding bias |
| --- | --- | --- | --- |
| Ahola 2010 [47] | + | + | - |
| Ahola 2012 [48] | + | - | - |
| Cohen 2004 [44] | - | + | - |
| He 2006 [42] | + | + | - |
| Kordbagheri 2024 [46] | - | + | - |
| Koponen 2017 [41] | + | + | - |
| Koponen 2019 [21] | + | + | + |
| Lei 2018 [45] | + | + | - |
| Lin 2009 [18] | ? | + | - |
| Márquez-Palacios 2020 [22] | - | + | - |
| Odajima 2018 [20] | - | + | + |
| Renosky 2010 [43] | + | + | - |
| Shiu 2004 [19] | - | + | - |
| Vega-Martínez, López-Martínez and Del-Pino-Casado [49] | - | + | + |

Notes: (+) low risk of bias; (-) high risk of bias; (?) There is not enough information for the assessment.

### Results of the review

Given that, on the one hand, a variety of results were found and many of them have hardly any studies, and on the other hand, some studies lacked adequate statistical data for meta-analysis, it was decided to first perform a narrative synthesis of the included studies and then a meta-analysis with the studies that presented adequate data for such meta-analysis.

### Narrative synthesis

The included studies can be classified into two types of outcomes: adherence to self-care and perception of the self-care plan.

#### Sense of coherence and adherence to self-care

Eleven studies [18,21,22,41,43–49] provided data on the relationship between the SOC and adherence to the self-care plan. Of these, seven studies showed a positive association (possible benefit) with adherence to the global self-care plan [18,21,22,43–46]; with adherence to the diet, four studies [21,41,48,49]; with adherence to recommendations on physical activity, two studies [41,48] and with medication adherence, one study [49]. On the other hand, in one study [47], no statistical association was found between the SOC and the follow-up of visits to the nurse and the physician.

#### Sense of coherence and perception of the self-care plan

Three studies [19,20,42] reported findings on the relationship between the SOC and the perception of the self-care plan. All three studies found an inverse relationship (possible benefit) between SOC and diabetes-specific stress [42], burden by self-care plan [20] and fear of hypoglycemia [19].

### Meta-analysis

Seven studies [18,21,22,43–46] provided data to quantitatively analyze the relationship between SOC and self-care adherence, finding a positive and moderate association (r= 0.32; 95% confidence interval (CI): 0.29, 0.35; N= 3,985; Average per study: 569.3), i.e., high SOC is associated by greater self-care adherence. All seven studies have positive statistical associations, and all the confidence intervals of the correlation coefficients of each study overlap at the same point (Fig 2).

**Fig 2.**
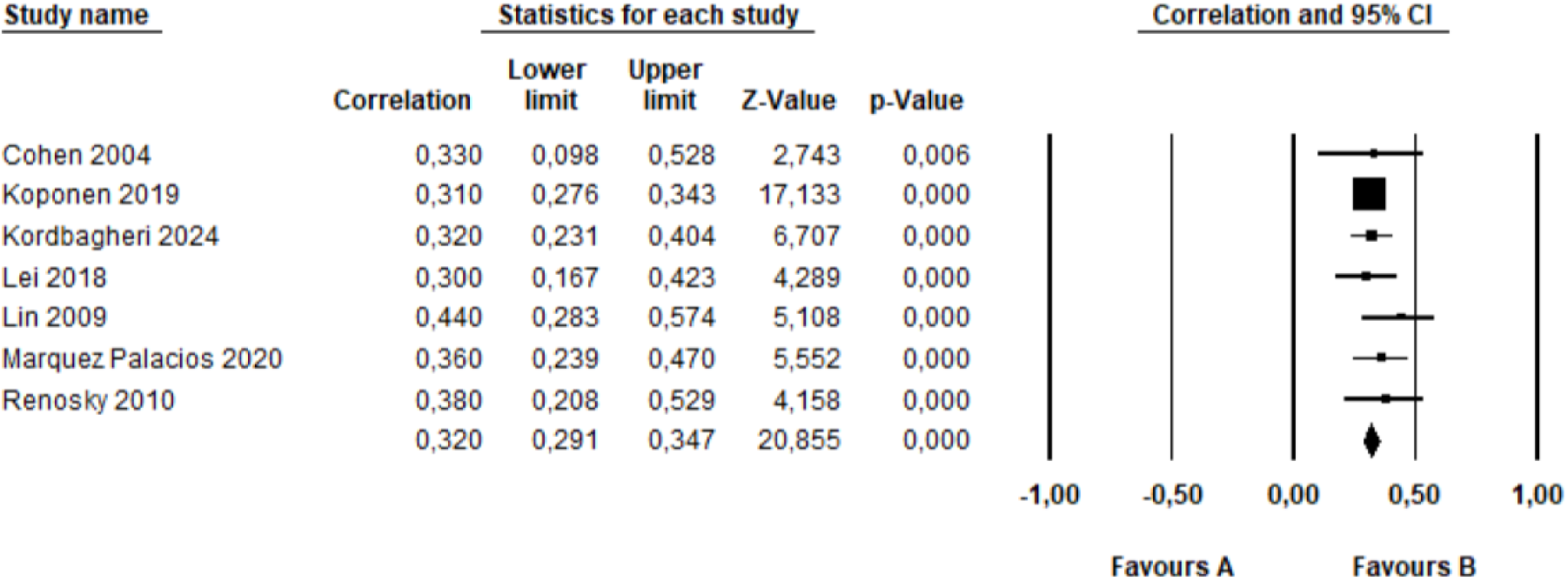
Forest plot for sense of coherence al adherence to self-cares.

There was no heterogeneity between the results of the different studies (Q= 3.7, degrees of freedom: 6, p= 0.72; I^2^ = 0.0). In addition, the prediction interval (true value in 95% of comparable populations), which is: 0.27, 0.37, is very similar to the confidence interval, demonstrating the absence of total heterogeneity.

Regarding publication bias, the funnel plot (Fig 3) appears asymmetric, although no effect is observed from small studies, and the Trim and Fill-adjusted value of the combined effect is 0.31 (3.1 % of variation). Therefore, it seems that there is publication bias but this bias has little effect on the result of the meta-analysis. The Egger’s test has not been performed because the number of included studies is less than 10 [50].

**Fig 3.**
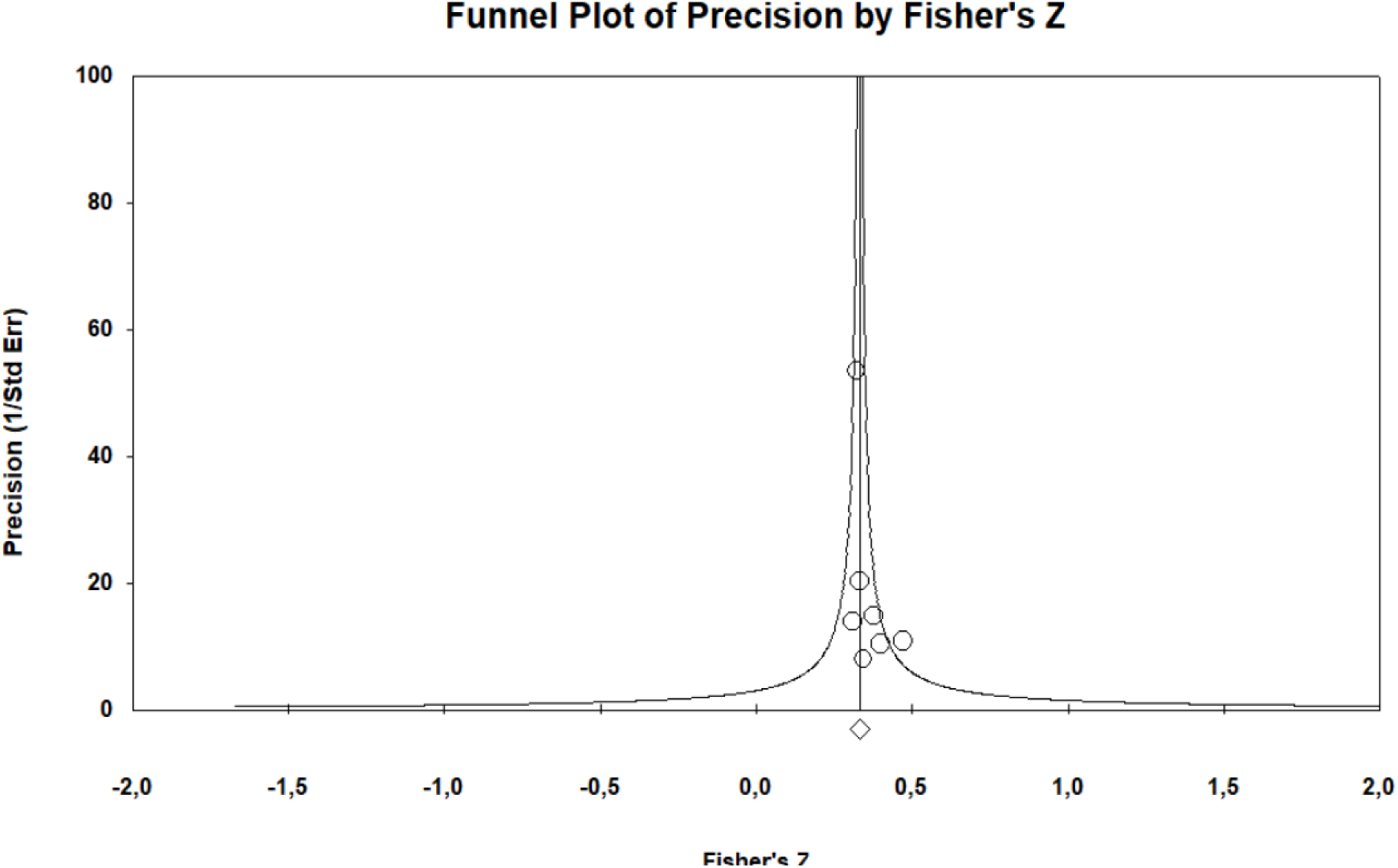
Funnel plot for sense of coherence al adherence to self-cares.

Regarding the sensitivity analysis, when performing different meta-analyses eliminating one study at a time, we found that the maximum variation of the combined effect is 7.5%, revealing an acceptably robust result.

Regarding subgroup analyses, they were performed for the type of DM, selection bias and confounding bias, since all studies included in the meta-analysis were descriptive cross-sectional and had a low risk of classification bias. No differences were found between studies that included people with T1D and T2D (r= 0.36; 95% CI: 0.23, 0.48; N= 479; 2 samples) and those that included only people with T2D (r= 0.32; 95% CI: 0.29, 0.35; N= 479; 4 samples). No differences were found between studies with probabilistic (r= 0.31 95% CI: 0.28, 0.34; N= 479; 3 samples) and non-probabilistic (r= 0.38; CI 0.29, 0.46; N= 479; 3 samples) samples, nor between studies that presented confounding bias control (r= 0.31; 95% CI: 0.28, 0.34; N= 479; 1 sample) and studies that did not (r= 0.36; 95% CI: 0.29, 0.42; N= 479; 5 samples).

## Discussion

The present study analyzed the relationships between SOC and the self-care management in people with T1D or T2D, showing that a high SOC is accompanied by better adherence to self-care and more positive perceptions of performing these self-care activities. To our knowledge, our meta-analysis on SOC and self-care adherence in people with DM is the first to be carried out worldwide.

The narrative analysis showed a positive relationship between SOC and adherence to diet, physical activity and medication. No relationship has been observed between SOC and the follow-up of visits by health professionals, although we only found a single study that analyzes this variable, so it would be advisable to carry out further research to provide information on this possible relationship. On the other hand, it is observed that high SOC could have beneficial effects in terms of the perception of the self-care plan. These results are consistent with the findings of Ludman and Norberg [51], who point to SOC as a factor contributing to emotional health and better adjustment to insulin treatment in people with DM. Since the positive perception of the self-care plan is related to better adherence to it, the SOC could improve adherence by eliciting positive perceptions regarding the self-care plan [52,53].

Our meta-analysis has robust results, no heterogeneity and the possible publication bias does not seem to affect these results. Regarding precision, the average number of participants per study and the number of studies included in the meta-analysis give us moderate precision.

Our results are in line with a study carried out in people with T1D on the relationship between SOC and clinical risk factors, which shows that people with high SOC have lower LDL cholesterol levels and therefore, that SOC could be a protective factor for cardiovascular complications [54]. Regarding research in populations with other chronic diseases, we find the work carried out by Moya [55], who analyzed the relationship between SOC and adherence to antiretroviral treatment in HIV-positive people and found a moderate and positive relationship. These results are consistent with research carried out in women with breast cancer, highlighting the relationship between high SOC and adherence to treatment [56]. It is also worth noting a review carried out in people with chronic diseases [53] that highlighted the relationship between SOC and a better perception of the disease and a lower risk of depression. Regarding cardiovascular disease, a better capacity for self-care was found in those with a high SOC and, on the contrary, those with a low SOC was linked to unhealthy behaviors. Finally, a study conducted in people with prediabetes revealed the relationship between high SOC and healthier behaviors in relation to diet, physical exercise and exposure to tobacco [57].

Antonovsky [58] explains these events by referring to the fact that adversities in life are better coped by people with high SOC. Stressful events are seen as meaningful, understandable, predictable, and manageable, so they are perceived as less threatening and therefore less stressful. For Antonovsky, people with high SOC are more likely to adopt healthy behaviors and have a more stable of self-reported level health in the face of stressful life events, compared to those who have a low SOC.

Therefore, SOC may be a tool in Health Promotion that could be useful in clinical practice and help professionals to anticipate and foresee non-adherence to the care plan of a person with DM with low SOC. Within this framework, more intense interventions could be carried out in these people, including, among other things, individual and group health education, with the aim of increasing both the SOC and other aspects that improve the ability to manage the care plan. It would be interesting to monitor these interventions for the future to establish a feasible salutogenic health education program and extend the salutogenic view through our health system. For this purpose, it is essential to carry out more studies such as the one conducted by Thompson [59] highlighting the effects of interventions to strengthen SOC in vulnerable people, or the review by Langeland et als. [60], who presents a range of different interventions carried out in various populations for the same purpose.

### Strength and limitations

Our study had several limitations. First, all the included studies are cross-sectional, therefore we cannot stablish causality. Second, several of the included studies had high risk of selection bias and confounding bias, although subgroup analyses did not show differences between studies with and without both biases. Third, people with T1D included in the studies is considerably smaller compared to the percentage of people with T2D included, leading to potential underrepresentation of this group. However, subgroup analyses did not reveal differences between studies with T1D and T2D diabetes and those with T2D.

### Recommendations for Further Research

Longitudinal studies that present temporal sequence are necessary in order to establish causal relationships between the variables studied and greater certainty of the involvement of SOC in self-care.

### Implications for policy and practice

Considering the level of SOC in the implementation of the self-care plan may lead to better rates of adherence to self-care and therefore improve the evolution of diabetes, reduce macro and microvascular consequences and mortality.

## Conclusions

Despite these limitations, we conclude that SOC may be a protective factor against non-follow-up of self-care in people with T1D and T2D. Longitudinal studies are necessary to confirm this protective effect.

## Availability of Data

The data that support the findings of this study are available from the corresponding author, [CLM], upon reasonable request.

## Acknowledgments

There are no thanks to be made.

## Author contributions

Vega-Martínez, María del Carmen: Conceptualization, Investigation, Validation, Data curation, Formal analysis, Visualization, Project administration, Writing – original draft, Writing – review & editing.

López-Martínez, Catalina: Validation, Investigation, Visualization, Supervision, Writing – original draft, Writing – review & editing.

Del-Pino-Casado, Rafael: Conceptualization, Methodology, Software, Data curation, Formal analysis, Investigation, Validation, Resources, Visualization, Supervision, Project administration, Writing – original draft, Writing – review & editing.

All authors have approved the final article.

