## Supplementary material for "Sense of coherence and self-care in people with diabetes: systematic review": S1 Appendix

**Full title**

[1]Department of Nursing, University of the Balearic Islands. Palma de Mallorca. Balearic Islands. Spain

Department of Nursing, University of Jaén. Jaén. Andalusia. Spain.

* **Corresponding author**

**Key words**

Diabetes mellitus; Self-care; Sense of coherence; Systematic review; Meta-analysis.

**S1 APPENDIX.**

**SEARCH STRING IN EACH DATABASE**

- PUBMED

Diabet* AND ("sense of coherence" OR Salutogenesis) AND (adherence OR compliance OR diet OR "physical activity" OR exercise OR insulin OR medication OR treatment* OR drug therapy OR self-management OR self care OR self-care OR self-control OR care plan* OR care management)

- CINAHL:

Diabetes mellitus AND (“sense of coherence” OR Salutogenesis) AND AB ((adherence OR compliance OR diet OR "physical activity" OR exercise OR insulin OR medication OR treatment* OR drug therapy OR self care OR self-management OR self control OR care plan* OR care management))

- PSYCINFO

Diabetes AND ("sense of coherence" OR salutogenesis) AND AB (adherence OR compliance OR diet OR "physical activity" OR exercise OR insulin OR medication OR treatment* OR drug therapy OR self-management OR self care OR self control OR care plan* OR care management)

- SCOPUS

TITLE-ABS-KEY(diabetes) AND TITLE-ABS-KEY("sense of coherence" OR salutogenesis) AND (TITLE-ABS-KEY(adherence) OR TITLE-ABS-KEY (compliance) OR TITLE-ABS-KEY (diet) OR TITLE-ABS-KEY ("physical activity") OR TITLE-ABS-KEY (exercise) OR TITLE-ABS-KEY (insulin) OR TITLE-ABS-KEY (medication) OR TITLE-ABS-KEY (treatment) OR TITLE-ABS-KEY (drug therapy) OR TITLE-ABS-KEY (self care) OR TITLE-ABS-KEY (self-management) OR TITLE-ABS-KEY (self control) OR TITLE-ABS-KEY (care plan*) OR TITLE-ABS-KEY (care management))
